# Shifts in Indonesia’s malaria landscape: an analysis of 2010-2019 routine surveillance data

**DOI:** 10.1101/2024.08.25.24312562

**Authors:** Bimandra A Djaafara, Ellie Sherrard-Smith, Thomas S Churcher, Sri Budi Fajariyani, Helen Dewi Prameswari, Herdiana Herdiana, Riskha Tiara Puspadewi, Karina D Lestari, Iqbal RF Elyazar, Patrick GT Walker

## Abstract

**Background:** Indonesia faces challenges in achieving its goal of eliminating malaria by 2030, with cases stagnating between 2015 and 2019 despite a decline in 2013. This study aims to analyse epidemiological trends and demographic changes in malaria cases regionally from 2010 to 2019, considering differences in surveillance across the country.

**Methods:** We used national and sub-national malaria routine surveillance data, applying statistical models to derive insights for future interventions. The analysis focused on metrics such as malaria incidence, test positivity, case demographics, and parasite species.

**Findings:** We estimate a progressive decline in malaria incidence in six of seven Indonesian regions over the study period, adjusting for increased testing from 2015 onwards. In these regions, cases have shifted to older, predominantly male demographics, suggesting a move from household-based to occupational transmission. However, in Papua, cases remain high and concentrated in children. Although Papua comprises just 2% of Indonesia’s population, its share of national malaria cases increased significantly from 40% to 90% between 2010 to 2019.

**Interpretation:** Since 2010, malaria trajectories in Indonesia have diverged, influencing sub-national control priorities. In most regions, progress towards elimination requires targeted interventions for high-risk populations and inter-district cooperation. In contrast, Papua struggles with high transmission rates despite mass insecticide-treated net campaigns. Achieving similar progress in Papua as in other regions is challenging yet crucial. Therefore, Papua could be a prime candidate for enhanced malaria management, maximising efforts towards larval source reduction, education, chemopreventive intervention, and vaccine.

**Funding:** Medical Research Council and Wellcome Africa Asia Programme Vietnam.

**Research in context:** *Evidence before this study:* As the country with the fourth biggest malaria burden outside of Africa, malaria control and elimination have been significant public health priorities in Indonesia, especially since the National Ministerial Decree on Malaria Elimination was passed in 2009. To understand the existing literature on malaria in Indonesia, we conducted a literature search on PubMed using the keywords ’MALARIA’ and ’INDONESIA’ for titles and abstracts from 1 January 2008 to 3 January 2024, yielding 386 results. Among these, only a few studies extensively discussed the broad landscape of malaria in Indonesia. Notably, Elyazar et al., in their series of studies published in the early 2010s, provided an in-depth look at the epidemiology of malaria in Indonesia, the history of malaria control efforts leading up to the 2009 decree, potential challenges in malaria control, and estimates of malaria prevalence across the country at the beginning of the 2010 decade. These studies laid a foundational understanding of the malaria situation in Indonesia at the start of the decade, capturing the diverse and complex nature of the elimination challenge. Sitohang et al. wrote a commentary article on the accelerated progress towards malaria elimination in Indonesia from 2007 to 2017. However, they also highlighted the persisting challenges that Indonesia faces in meeting the 2030 elimination target. Therefore, there remains a need to quantitatively assess the impact of acceleration strategies ten years after the 2009 ministerial decree and understand the shifting epidemiological patterns of malaria, especially in the context of Indonesia’s diverse and changing demographic and environmental landscapes.

*Added value of this study:* Our study offers a significant contribution to understanding contemporary malaria epidemiology in Indonesia, especially after a period of acceleration in malaria elimination efforts. We did a comprehensive analysis of a decade’s worth of malaria surveillance data in Indonesia, revealing diverging trends in malaria incidence between regions and the changing profiles of malaria cases. We highlight the significant decline in malaria cases since 2010 in six out of seven regions in Indonesia, with cases shifted to older and predominantly male demographics, indicative of a move from household-based to occupational transmission. However, in Papua, cases remain high and concentrated in children despite efforts such as mass insecticide-treated nets distribution campaigns. In 2019, Papua contributed to 90% of malaria cases across the country, an increase from around 40% in the early 2010s despite comprising only 2% of Indonesia’s population.

*Implications of all the available evidence:* This study identifies challenges and discusses the way forward for Indonesia’s fight against malaria. Although there has been great progress, the findings suggest that regionally tailored strategies are needed for effective elimination efforts going forward. In areas where malaria incidence has declined and the demographics of typical malaria cases have shifted, malaria interventions should be focused on the high-risk population in respective regions, which includes mobile and migrant populations such as forest workers and indigenous people, amongst others. Furthermore, inter-district cooperation is needed to prevent malaria importations and reintroductions to districts already eliminating or achieving progress towards malaria elimination. In Papua, where malaria transmission remains high, novel and innovative interventions may be required to accelerate progress towards malaria elimination. While some malaria vaccines have proven effective in high-burden countries within Africa, severe disease numbers are lower in Papua, and sustained coverage levels with routine immunisations have been a challenge to maintain. Hence, Indonesia is hesitant to adopt such a strategy. Additionally, Indonesia also has challenges in controlling *P. vivax*, which presents a significant burden on the population and is a problem not seen in African countries. Therefore, in Indonesia, chemopreventive interventions (such as intermittent preventive treatment of malaria during pregnancy or IPTp), and additional vector control interventions (such as larval source reductions) may be potential tools to deliver progress for Papua and other remaining high-burden locales. Furthermore, vaccine development efforts to target adults and *P. vivax* may also be useful additions for controlling malaria in Indonesia in the future.

## Introductions

Designing optimal malaria control strategies in Indonesia is a substantial challenge due to a multitude of complex epidemiological factors.^1^ The country’s diverse landscape of endemicity, population densities that range from dense urban areas to sparsely populated rural regions, a variety of malaria vectors with differing behaviours and bionomics, and the co-endemic presence of two dominant malaria species, *Plasmodium falciparum* and *Plasmodium vivax*, all contribute to this complexity.^1,2^ Additionally, concerns around zoonotic malaria parasite (*Plasmodium knowlesi*) infections^3^ and the challenges of controlling malaria in mobile and migrant populations, particularly in areas nearing elimination,^4^ further complicate the situation.

Despite these challenges, from 2010-2019, Indonesia’s malaria elimination efforts made substantial progress. In 2017, more than half of the country’s districts—accounting for roughly 72% of the population—reported no local malaria transmission for three consecutive years, marking them malaria-free.^5^ This achievement is largely attributed to the intensification of control efforts from the early 2000s and a National Ministerial Decree on Malaria Elimination in 2009,^6^ which granted local authorities the autonomy and political backing to implement effective, locally tailored malaria control. The decree also precipitated improvements in aspects such as financing, the scaling up of artemisinin combination therapy (ACT), mass distributions of long-lasting pyrethroid-insecticide treated nets (LLINs), mandated laboratory confirmations, quality assurance for diagnoses, screening and treatment for pregnant women, and enhanced surveillance and reporting.^5,7^

After roughly ten years following the decree, it is crucial to objectively and quantitatively measure the impact of these endeavours to ensure the effective and efficient deployment of future control and elimination efforts. Routine surveillance data for malaria have increasingly been utilised to set national and regional targets, estimate disease burden, and measure the impact of control strategies.^8^ In Indonesia, malaria case surveillance and reporting coverage improved significantly from covering only 26-50% of districts in 2010 to over 75% in 2015.^5^ However, these achievements in surveillance strengthening provide a challenge to interpreting true temporal patterns of underlying case trends within the reported data.

In this study, we leveraged a decade of routine malaria surveillance data from the Indonesia National Malaria Control Program (NMCP) to better understand the progress in malaria control and elimination efforts across the diverse Indonesian landscape, developing an inferential framework to adjust reported trends for the changes in surveillance capacity that occurred during the period. We characterise the overall trends in metrics such as case counts and test positivity ratios (TPR) and examine the shifts in malaria case profiles, including proportions by parasite species, and the age and sex of cases. We then compared their patterns to those that have been observed in other settings as they move toward malaria elimination, such as *P. vivax* becoming the dominant malaria species,^9–11^ an increase in the average age of clinical disease,^12,13^ and an increase in the proportion of cases in men, as exposure becomes more occupational-driven.^9,14^ Through this multi-faceted lens, we aimed to uncover a more nuanced understanding of the trends of the ‘true’ burden of malaria in Indonesia after a decade of increased malaria control efforts.

## Methods

### Indonesia routine malaria surveillance data

We utilised monthly aggregated district-level malaria routine surveillance data from the SISMAL (*Sistem Informasi Surveilans Malaria* or Malaria Surveillance and Information System) platform of Indonesia NMCP from 2010 to 2019.^15^ For this study, monthly aggregated malaria cases and tests data were used. The total number of malaria diagnostic tests data are a combination of tests performed using either microscopy or rapid diagnostic test (RDT). The aggregated case data were grouped by age, sex, and parasite species. The age groups comprise 0-4 years old, 5-9 years old, 10-14 years old, and ≥ 15 years old. The parasite species recorded were *P. falciparum*, *P. vivax*, *P. knowlesi*, *Plasmodium malariae*, *Plasmodium ovale*, and mixed infections of *P. falciparum* and *P. vivax*. However, the aggregated data did not cross-tabulate malaria cases across age groups or sex with parasite species.

### Estimating trends of malaria metrics from routine surveillance data

We used Generalised Additive Models (GAMs) to produce estimates of overall regional trends adjusted for reporting at the district level throughout 2010-2019, while capturing complex non-linear trends between covariates and response variables using smooth functions or splines.^16,17^ Each region of Indonesia (Sumatra, Java & Bali, Kalimantan, Nusa Tenggara, Sulawesi, Maluku, and Papua) had a model fitted independently for each malaria metric, incorporating district and month-year as random effects covariate and a penalised smoothing spline covariate, respectively.

We employed GAM with a negative binomial family, to account for likely overdispersion in the distributions of cases and tests, and population counts as the model offset variable to model malaria cases and tests per 1,000 population. Metrics and case profiles measured as proportions were modelled using GAMs with a binomial family and logistic link function. The mgcv package in R^18^ was used for the implementation of GAM.

The malaria surveillance metrics and case profiles modelled are shown in **Table 1**, alongside their respective distribution families. To estimate the regional-level trend lines for all metrics, we calculated the weighted average values of all district-level trend lines within a region. The weighting factors used were: 1) district-level population counts for modelled cases and tests; 2) modelled district-level tests for TPR; and 3) a combination of modelled district-level TPR multiplied by modelled tests for proportions of *P. vivax* cases, cases in males, and cases in ≥ 15 years old.

**Table 1.** The malaria surveillance metrics and cases profiles modelled and their respective distribution families.

| Malaria surveillance metrics and case profiles | Distribution family |
| --- | --- |
| Cases per 1,000 population (overall and by age groups) | Negative binomial |
| Tests per 1,000 population | Negative binomial |
| Test positivity ratio (TPR, %) | Binomial (logit) |
| Proportion of $\geq 15$ years old cases (%) | Binomial (logit) |
| Proportion of cases in males (%) | Binomial (logit) |
| Proportion of *P. vivax* cases (%) | Binomial (logit) |

### Modelling the relationship between age of malaria cases and malaria endemicity

The previous section measured the relationship between the age of malaria cases (i.e., the proportion of cases in adults) and malaria endemicity indirectly by comparing trends of both metrics over time. Here, we developed a framework using a generalised linear model (GLM) to directly analyse the relationship between malaria endemicity and age to see whether consistent patterns were observed across all regions of Indonesia. We assumed a model whereby the Annual Parasite Incidence (API) per 1,000 (at the log-scale) alters the mean age of reported malaria cases (µ) on the geometric scale. The GLM was fitted to the proportion of malaria cases by age groups (0-4, 5-9, 10-14, and ≥ 15 years old). Each year’s district-level malaria case data were used for the model fitting process, filtering only observations with at least 30 malaria cases reported to reduce the noise from the low-level malaria case counts.

## Results

Figure 1 shows the national-level trends of several malaria metrics derived from the routine malaria surveillance data between 2010 and 2019. The number of reported malaria cases was reduced by half during this period, while malaria tests, which reflect surveillance efforts, almost doubled within the same period (Figure 1A). Nationwide, *P. falciparum* and *P. vivax* are the dominant malaria parasite species, with *P. falciparum* reported more frequently throughout the decade (Figure 1B). Malaria cases per capita were found more frequently in males (Figure 1C) and the youngest age groups (Figure 1D). However, those youngest age groups also experienced the largest declines in the per capita malaria rate over the decade.

**Figure 1.**
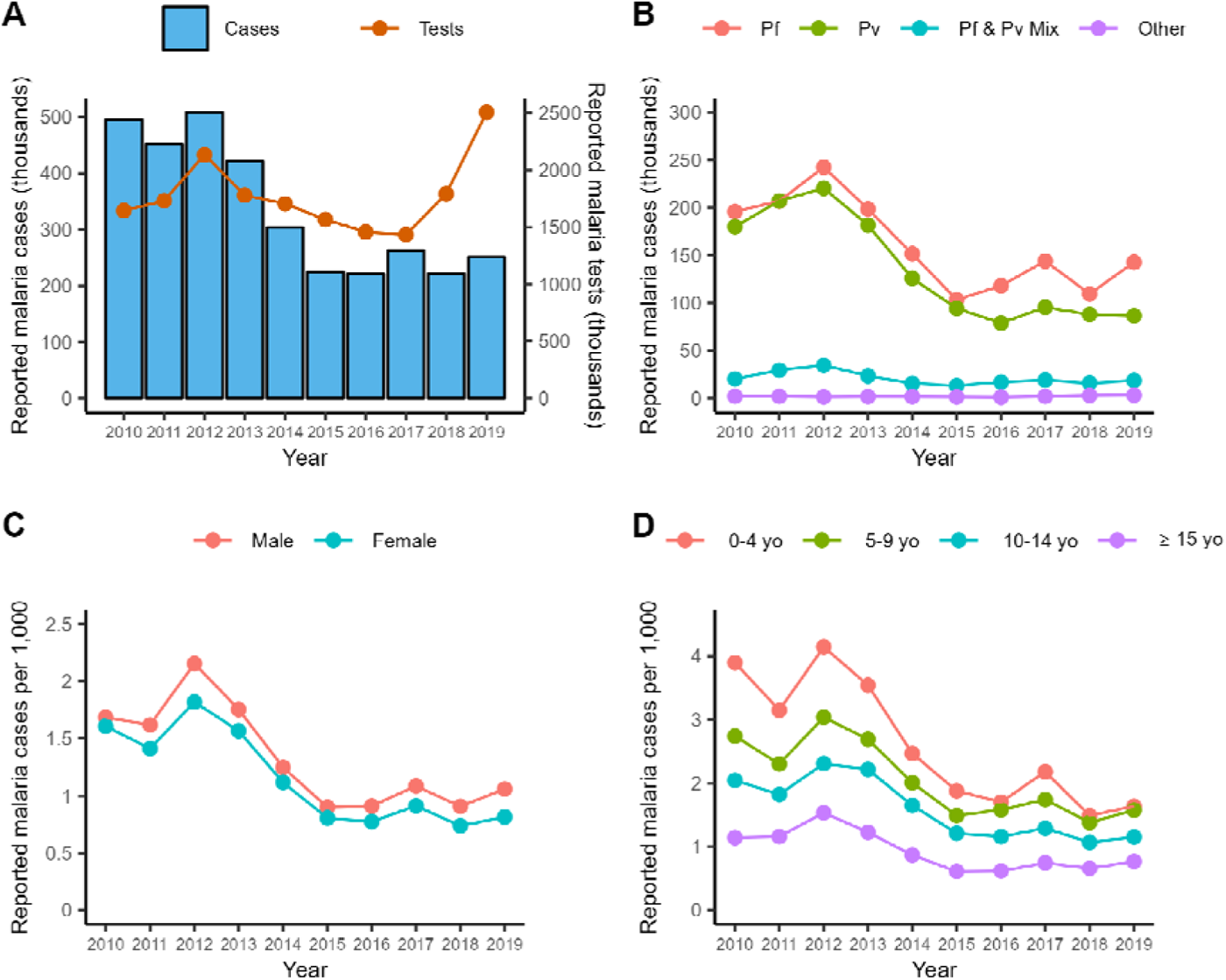
National-level trends of several malaria metrics, calculated yearly, derived from routine malaria surveillance data. A) reported malaria cases and tests performed; B) reported malaria cases by parasite species; C) reported malaria cases by sex; and D) reported malaria cases by age groups.

The geographical heterogeneity of malaria transmission in Indonesia is shown in Figure 2. In most regions, both median and maximum district-level malaria endemicity and maximum district-level incidence fell steadily throughout the study period (Figure 2A). However, some districts, particularly in the Papua region (the easternmost region), showed considerable differences in trends. Here, despite overall showing a progressive reduction in median district-level malaria endemicity, some districts reported similar or higher API per 1,000 in 2019 relative to 2010 (Figure 2B).

**Figure 2.**
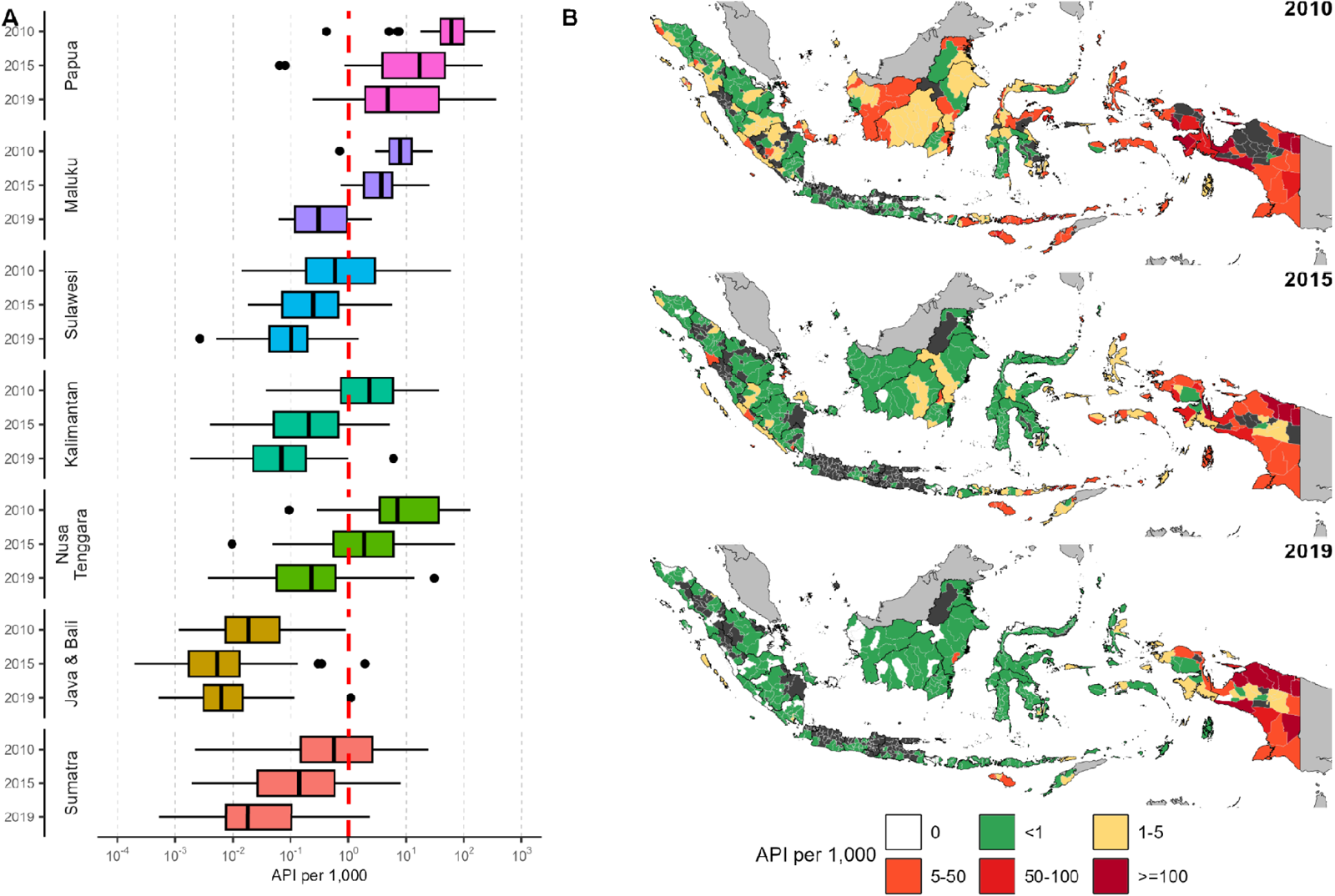
Distributions of malaria incidence in Indonesia. A) Within-region boxplots of API per 1,000 in 2010, 2015, and 2019 (from the top to the bottom row, respectively); and B) Geographical distributions of API per 1,000 at the district-level in 2010, 2015, and 2019. Dark greys denote no data was available.

Figure 3 shows the region-level trends of several malaria metrics estimated by GAM between 2010 and 2019, adjusting for reporting rates by district across the study period. Over the decade, adjusted malaria incidence rates declined in all regions of the country (Figure 3A). The declining trends differ from one region to another in terms of their baselines and slopes. Hence, there are differences in the magnitudes of the decline, with the highest reduction magnitude estimated in the Sumatra region (81·5-fold reduction) and the lowest in the Papua region (3·6-fold). Cases per 1,000 population trends for each age group are shown in **Figure S1**. These declines in malaria cases per 1,000 population were estimated despite case-finding efforts showing no declining trend across all regions (Figure 3B). The estimated trends of testing efforts are shown on a per 1,000 population basis, meaning that, in absolute terms, as the population grew over the years, the testing efforts increased. The substantial reduction in malaria burden across all Indonesian regions is also supported by the estimated test positivity ratio (TPR) trends, which show declines across all regions (Figure 3C). However, in the Papua region, we observed an increase in adjusted TPR in 2017 before declining again in the following years.

**Figure 3.**
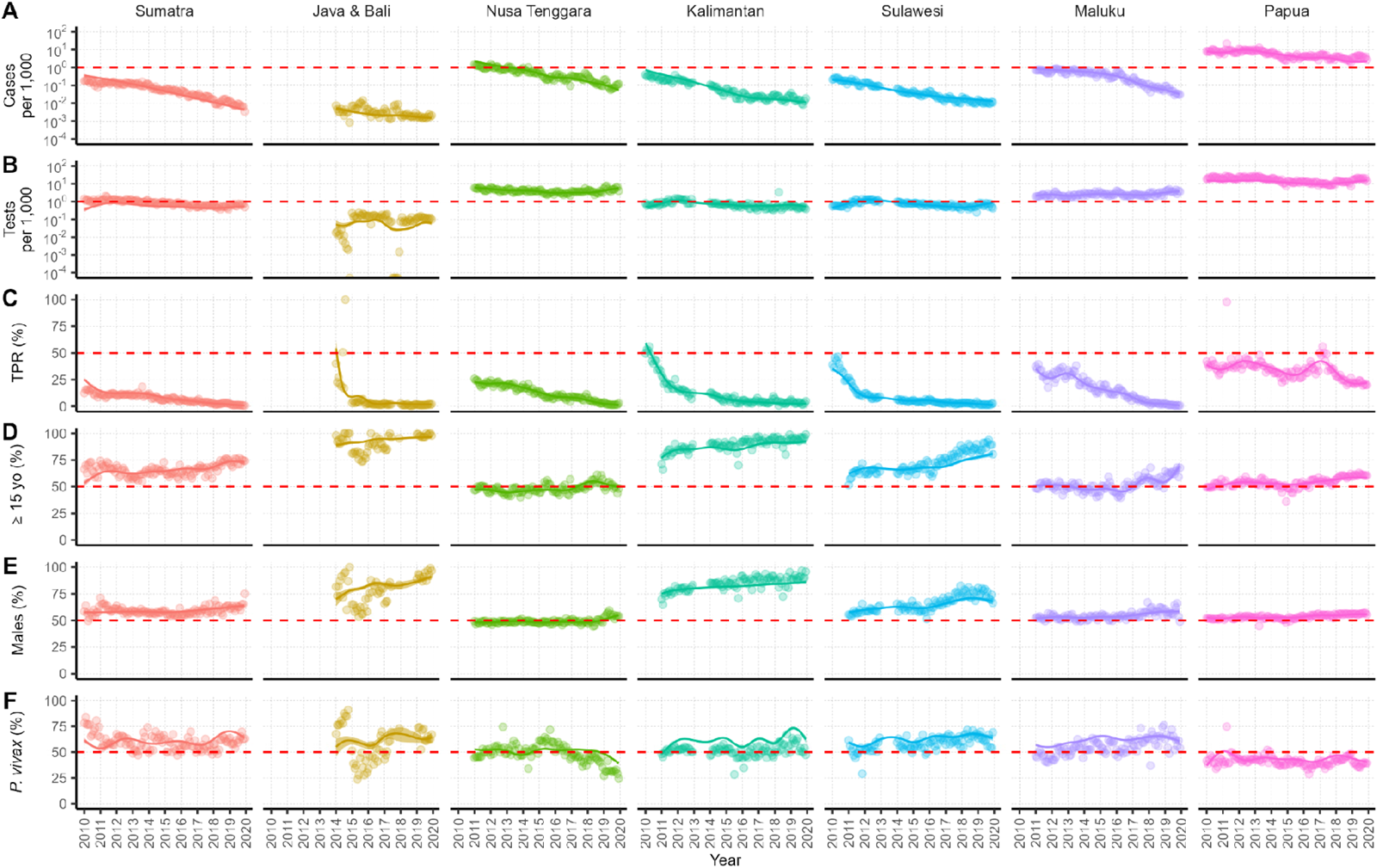
Regional-level monthly trends of several malaria metrics derived from routine malaria surveillance data. Solid lines and the shaded areas denote the median and 95% credible intervals of the modelled trends using GAM. The semi-transparent points denote region-level monthly averages from data. The metrics shown are: A) cases per 1,000; B) tests per 1,000; C) TPR (%); D) proportion of cases in ≥ 15 years old age group (%); E) proportion of cases in males (%); and F) proportion of P. vivax cases (%). Red dashed line is included as a fixed value to aid comparison between areas.

Cases were generally found in older populations, with increasing proportions of malaria cases in adults over the years (Figure 3D). However, on a per capita basis, malaria burden in children is still the highest (Figure 1D). Furthermore, malaria cases have become increasingly male-dominant (Figure 3E), which could indicate a shift towards a higher proportion of occupational exposures. In terms of malaria parasite species, there is no sign that *P. vivax* became the largely dominant parasite species in any region (Figure 3F), despite some regions experiencing slight shifts in species distribution. Notably, Nusa Tenggara is the only region where there has been a consistent decline in the proportion of *P. vivax* infections in the last years of the decade. Papua, on the other hand, is the only region with consistent *P. falciparum*-dominant infections in the country.

The relationship between the different adjusted metric trends was estimated using Spearman’s rank correlation, combining model estimates from all regions and for each region (Figure 4). There is a strong positive correlation between malaria cases and TPR at both national and regional level, though somewhat less so in Papua region. Meanwhile, decreases in both cases and TPR showed correlation with increases in the proportion of cases that were adult, which itself was largely synchronised with increases in the proportion of cases that were male. This trend of cases becoming typically older and male as transmission declines was particularly strong in regions of historically lower endemicity (Java & Bali, Sumatra, Sulawesi, Kalimantan). National-level increases in the proportion of cases that were *P. falciparum* (Figure 1B) mask high regional-level correlations between declines in transmission and the increasing role of *P. vivax* in all but the two regions with the highest burden (Nusa Tenggara and Papua).

**Figure 4.**
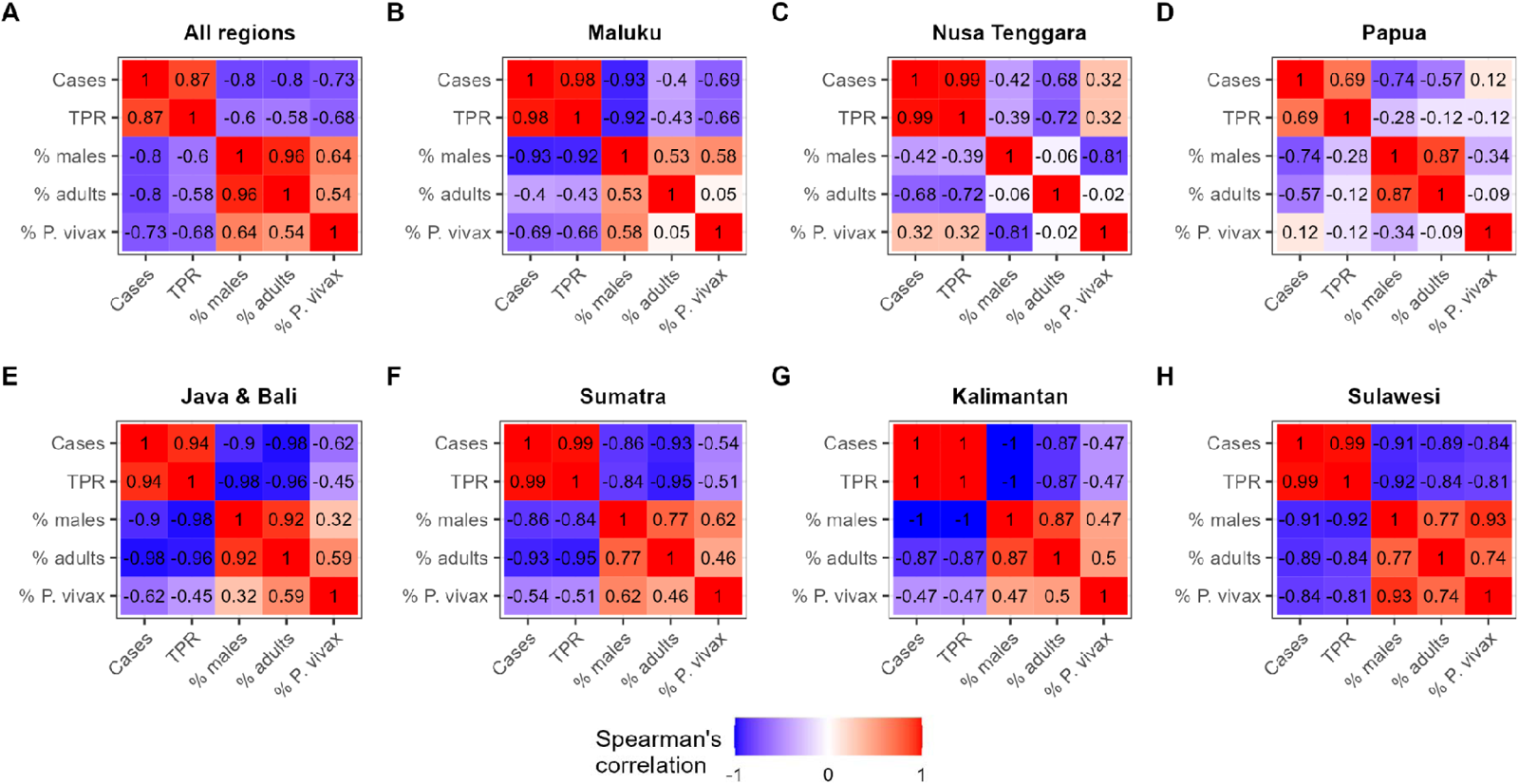
Spearman’s rank correlations between the modelled monthly estimates of malaria metrics using GAM. Red denotes positive correlations, while blue denotes negative correlations between metrics. A) All regions; B), C) & D) Regions with the highest malaria endemicity: Maluku, Nusa Tenggara, and Papua, respectively; E), F), G) & H) Regions with the lowest malaria endemicity: Java & Bali, Sumatra, Kalimantan, and Sulawesi.

Finally, we investigated how malaria endemicity (as measured by API per 1,000) shapes the age-profile of reported cases using GLM. Figure 5 illustrates the relationship between API per 1,000 and the proportion of cases in the population aged 0-4, 5-9, 10-14, and ≥ 15 years old. In low-endemic settings (for example, Java & Bali), cases are dominated by adults, but the proportion of cases in children increases as endemicity increases. Model parameters convergence and validation, as well as proportion of cases by age group (in selected districts representing the upper, middle, and lower quantiles of API) are shown in **Figure S2**, **S3**, and **S4**. **Figure S5** shows the combined modelled relationship between API per 1,000 and the proportions of those age groups.

**Figure 5.**
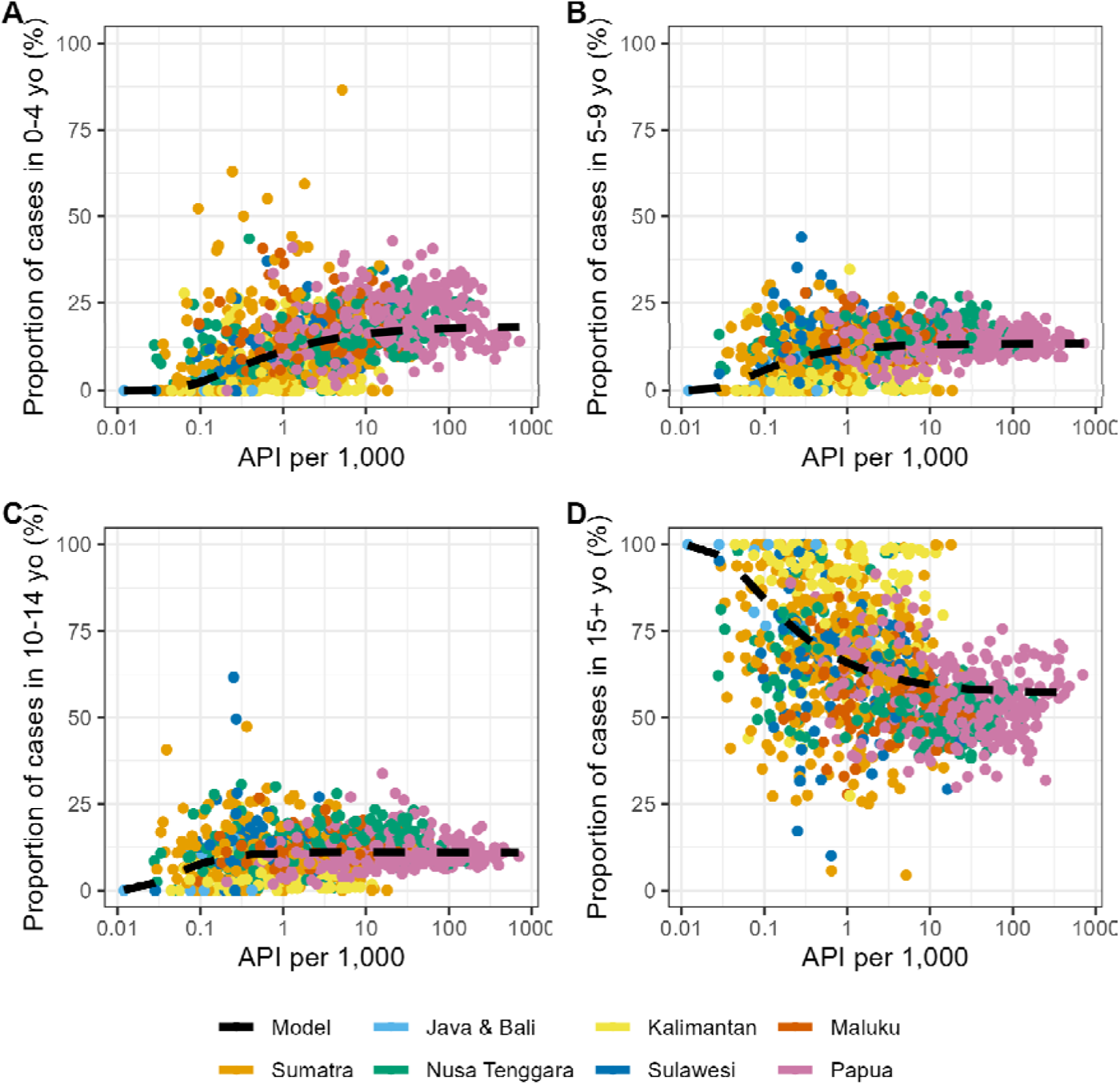
Modelled relationships between API per 1,000 and the proportion of cases in A) 0-4 years old; B) 5-9 years old; C) 10-14 years old; and D) ≥ 15 years old, with overlaid data from routine surveillance. Dashed lines denote the median of generalised linear model (GLM) estimates of the relationship. The colours of the data points represent regions.

Figure 6 shows the geographic distribution of outliers to the GLM results, whereby model estimates of the proportion of adults, generated using district-level case counts (Figure 6A) are compared to those observed in the data (Figure 6B). As Figure 6C shows, 84% of 400 districts reporting their malaria data in 2019 fall within the -20% to 20% difference bracket between data and model estimates, which interval arbitrarily chosen to visualise ‘no difference’ between them. Those that lie beyond this threshold include clusters of districts within Sumatra in 2019 that coincides with some of the steepest declines in API in the study period, where a higher proportion of children than expected appear in case-data than expected by the model. A similar pattern is also seen in low-endemic districts in western Kalimantan. This contrasts, however, eastern Kalimantan where districts often report higher than expected cases in adults throughout the study period.

**Figure 6.**
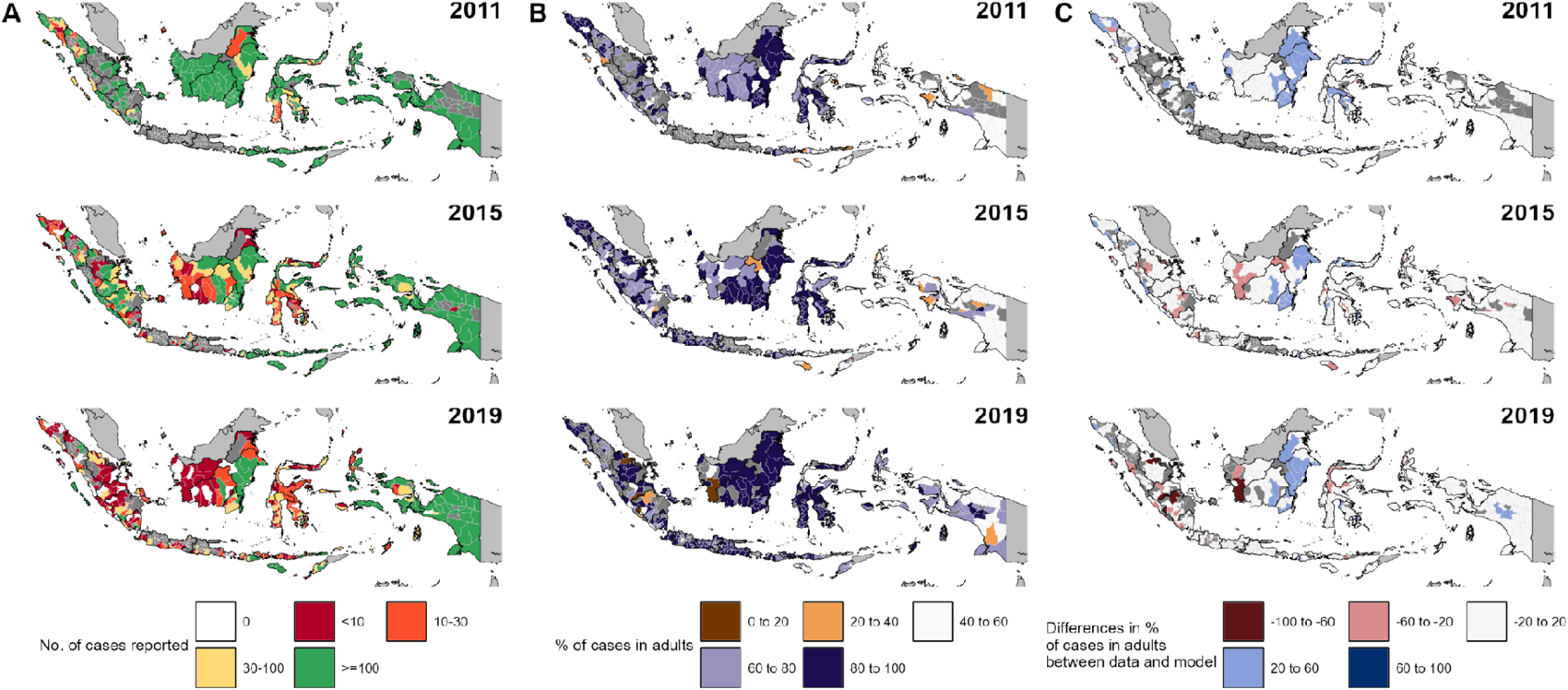
Maps highlighting the proportion of malaria cases in adults in Indonesia. The top, middle, and bottom rows represent 2011, 2015, and 2019, respectively. A) District-level maps of the number of malaria cases reported within a year, serving as ‘sample size’ of the calculated proportions from data. Districts reporting low case counts are coloured in red, representing low sample sizes to infer proportions presented in B), while districts reporting high case counts are coloured in green, representing high sample sizes; B) District-level maps of the reported proportion of malaria cases in adults. Brown colours denote districts where children dominated the reported malaria cases, while districts coloured in purple denote adult-dominant malaria cases; and C) District-level maps of the difference between the proportion of malaria cases in adults based on reported data and the model average, as shown in Figure 5. Red colours denote districts with lower proportions of cases in adults than the model average, while blue colours denote districts with proportions of cases in adults higher than the model average.

## Discussion

Our analysis highlights the heterogeneity in progress towards malaria elimination across Indonesia despite a major decline in malaria cases that occurred nationally between 2010 and 2019. While national-level malaria data would suggest stagnation in progress since 2015, sub-national trends tell a different story. In regions covering more than 95% of the country’s population, malaria cases have steadily decreased. Furthermore, while regions had differences in malaria transmission at baseline, progress towards malaria elimination in those regions also happened at different paces. Trends in raw national-level data have become increasingly dominated by high-endemic regions such as Nusa Tenggara, Maluku, and Papua, where only 7% of the population resides (19 million people) but which represented 95% of reported malaria cases in 2019, rising from 73% in 2010 (Papua region, 2% population, ∼40% contribution to ∼90%).

When data are considered at the region level, further divergence in trends between regions with lower and higher baseline endemicity emerges, with the four lowest endemicity regions (Java & Bali, Sumatra, Sulawesi, Kalimantan) showing clear patterns of a steadily rising proportion of cases in males and adults as transmission has declined. These findings are typically a sign of occupational-driven exposure, where transmission occurs in high malaria-risk settings such as forests and mines, far from human settlements.^4,19,20^ This suggests that control strategies in these regions may need to be reoriented to address these particular demographic groups more effectively.

Meanwhile, a concomitant shift in malaria species to *P. vivax* supports the need for effective approaches to achieving radical cure and elimination of the hypnozoite reservoir. A recent randomised control trials study showed that co-administration of single dose tafenoquine with dihydroartemisinin-piperaquine did not show a clinically meaningful benefit and thus does not support the combination as a radical cure for *P. vivax* in countries using ACT for *P. vivax* treatment,^21^ such as Indonesia. Short-course (7-day) regimen of primaquine might improve adherence and effectiveness of *P. vivax* radical cure, but it requires G6PD testing available at health facilities to ensure safety.^22^ Alternatively, community health centre-based strategies, such as supervised 14-day primaquine courses, may improve treatment adherence and lower *P. vivax* recurrence.^23,24^ However, addressing structural barriers hindering patient adherence is important to ensure their success.^25^

Our results also highlight the value of examining the relationship between surveillance metrics to identify districts where control measures may not be working as expected. When considering the relationship between case incidence and age of people with reported cases we found distinct patterns of younger than expected age-patterns emerging in clusters of districts in the northern province of Aceh, Sumatra and western provinces of Kalimantan. Many of these districts were those which had experienced some of the highest decline in malaria incidence over the past decade (i.e., from API per 1,000 > 100 to API per 1,000 < 10). This may implicate a role of residual immunity in adults^12^ and so this outlier status may prove transient. However, for some districts in the central region of Sumatra, deforestation due to increased mining and plantation activities has also increased malaria risks in the nomadic indigenous population, with malaria prevalence as high as 24%.^26^ Such pockets of community-based transmission, in a wider landscape of largely occupational exposure, would also contribute to the younger than typical demographics of observed cases in the region. This highlights the importance of protecting indigenous population from malaria to progress malaria elimination in this region. Meanwhile, we found that nationwide, in terms of having an older than expected age distribution for their given endemicity, the largest outliers were clustered within the remaining districts with API > 10 on Kalimantan, towards the east of the island. This region is well-known as an area of primarily occupational-driven exposure through agriculture and forest-related activities,^20,27–29^ highlighting the key ecological and economic issues that will need to be addressed in order for malaria elimination to be achievable in the region.

Maintaining progress toward elimination and optimising control in lower endemic regions remains an important consideration, yet the overwhelming and increasing percentage contribution of the Papua region to national case trends highlights this region as providing the largest obstacle to Indonesia’s elimination targets. In this region, case demographics indicate that a decade after the scale-up of vector control, transmission remains firmly embedded within the community and the household. Understanding ongoing barriers to progress in this region is key to ensuring progress at the national level. Still, such barriers are likely to vary at the local level, with districts in western Papua region showing sustained reductions whilst districts in the east of the region experienced resurgences in API in the latter part of the decade. An increase in cases reported was also partly due to the increase in the number of tests reported in the late 2010s. However, this resurgence also mirrored the trends observed in the neighbouring country Papua New Guinea.^30,31^ Further research is needed to explore whether the drivers of the resurgences in these regions were similar and how cross-border transmission contributes to risk.

Furthermore, the need for innovative and novel interventions in the Papua region is evident. Mass drug administration campaigns were conducted in several districts in Papua in 2023. The impact of these campaigns will be investigated. However, the expert committee in Indonesia has not recommended the current malaria vaccine (RTS and R21) to children in Papua because the level of severe disease is relatively low, and routine immunisation coverage in Papua is also low. The additional resources required for the malaria vaccine to be included within the routine immunisation programs would add an unreasonable burden to Essential Program on Immunisation (EPI) programs when it is known that the vaccine coverage will not be as high as expected and likely too low to achieve the same success as observed in African countries that are deploying the tool. It may be possible to include vaccine development efforts (including ones effective in adults and targeting *P. vivax*), other chemopreventive interventions (e.g. intermittent preventive treatment of malaria in pregnancy, IPTp^32^), and other vector control tools, such as larval source management, to strengthen existing control efforts aimed at rapidly reducing the malaria burden in Papua.

Trends in endemicity levels, case demographics and *Plasmodium* species composition provide less of a clear relationship in Nusa Tenggara and Maluku when aggregated at the regional level. This is likely to be in part due to the high levels of between-province and between-district heterogeneity within these regions. For example, in Nusa Tenggara the counter-intuitive increase in proportion of cases attributable to *P. vivax* in the context of a wider decrease in transmission is more readily explainable by region-level cases becoming increasingly concentrated in the southern archipelagic islands of Sumba and Timor (representing 86% of cases in Nusa Tenggara in 2019), where transmission has declined to a lesser extent than in other districts, and *Plasmodium* species composition remain largely unchanged. Meanwhile, for the remainder of the region (representing 75% of the total population), sustained declines in cases have coincided with a steady increase in the contribution of *P. vivax*, mirroring trends in other regions of Indonesia where cases have fallen to low levels.

Though better quantification of the heterogeneous landscape of malaria in Indonesia is crucial for improved malaria control, understanding the drivers of this heterogeneity is complex, particularly as the scaling up of malaria has coincided with many other ongoing climactic, environmental and societal trends occurring within the country. This includes changes in land-use due to agricultural expansion, deforestation, and other land-use modifications^33–35^ and variations in climate factors such as temperature, rainfall, and humidity across different regions of the country and climate variability may have contributed to the observed trends and regional differences.^36^ Finally, the capital relocation project from Jakarta to East Kalimantan might pose an additional risk of increased local transmission in the region due to the influx of malaria-naïve populations, the availability of malaria vectors, and its proximity to malaria-endemic districts.^37^ Further investigation into the interplay of these factors could provide a more nuanced understanding of the changes in malaria incidence observed throughout the decade.

Despite not examining the emergence of zoonotic malaria infections in the country, we also note the need for enhanced diagnostics and surveillance capacity. Past studies have shown how misdiagnosis was prevalent in areas with high risk of *P. knowlesi* infections.^38,39^ Furthermore, as *P. knowlesi* becomes the dominant malaria infection in neighbouring Malaysia Borneo,^40^ a recent survey in one of Indonesia’s bordering districts found that it is not the case yet in the Indonesia part of the island.^29^ More extensive surveillance is needed to confirm that *P. knowlesi* is currently not a threat to malaria elimination efforts in the country.

In conclusion, while significant progress has been made in the past decade towards malaria elimination in Indonesia, much work remains. Issues around malaria in mobile and migrant populations, changing environment and climate, zoonotic malaria, reducing *P. vivax* hypnozoite reservoirs, and, most importantly, malaria control in high transmission areas will be the main challenges as Indonesia works towards their malaria elimination goals. This study underscores the need for locally tailored interventions and greater inter-district cooperation to tackle malaria importation. These are crucial steps for Indonesia as it continues its ambitious goal of eliminating malaria.

## Data Availability

All data used in the present study are owned by the malaria working team at the Ministry of Health of the Republic of Indonesia. Requests for access to the relevant data may be made directly to the team.

## Contributors

BAD, ESS, TSC, IRFE, and PGTW conceived and designed the study. SBF, HDP, and RTP collected, verified, and provided data interpretation. BAD, ESS, TSC, and PGTW were involved in the data analysis and interpretation. BAD and PGTW drafted the paper. BAD, ESS, TSC, SBF, HDP, HH, RTP, KDL, IRFE, and PGTW critically revised the manuscript for important intellectual content, and all authors gave final approval for the version to be published.

## Disclaimer

The views in this article are those of the authors and do not necessarily represent the views, decisions, or policies of the institutions with which the authors are affiliated.

## Acknowledgements

BAD, ESS, TSC and PW acknowledge funding from the MRC Centre for Global Infectious Disease Analysis (reference MR/X020258/1), jointly funded by the UK Medical Research Council (MRC) and the UK Foreign, Commonwealth & Development Office (FCDO), under the MRC/FCDO Concordat agreement and is also part of the EDCTP2 programme supported by the European Union. ESS is funded by a UKRI Future Leaders Fellowship from the Medical Research Council (<u>MR/T041986/1</u>).

## References

1. Elyazar IRF, Hay SI, Baird JK. Chapter Two - Malaria Distribution, Prevalence, Drug Resistance and Control in Indonesia. In : Rollinson D, Hay SI, eds. Advances in Parasitology. Academic Press, 2011: 41–175.

2. Elyazar IRF, Sinka ME, Gething PW, et al. Chapter Three - The Distribution and Bionomics of Anopheles Malaria Vector Mosquitoes in Indonesia. In: Rollinson D, ed. Advances in Parasitology. Academic Press, 2013: 173–266.

3 Lempang MEP, Dewayanti FK, Syahrani L, et al. Primate malaria: An emerging challenge of zoonotic malaria in Indonesia. One Health 2022; 14: 100389.

4. Ekawati LL, Johnson KC, Jacobson JO, et al. Defining malaria risks among forest workers in Aceh, Indonesia: a formative assessment. Malar J 2020; 19: 441.

5 Sitohang V, Sariwati E, Fajariyani SB, et al. Malaria elimination in Indonesia: halfway there. Lancet Glob Health 2018; 6: e604–6.

6 Departemen Kesehatan. Keputusan Menteri Kesehatan Republik Indonesia Nomor 293/MENKES/SK/IV/2009 28 April 2009 tentang Eliminasi Malaria di Indonesia. Jakarta: Direktorat Pemberantasan Penyakit Bersumber Binatang, Departemen Kesehatan …, 2009.

7. The Global Fund. The Global Fund - Indonesia - Grant Implementation. 2024. https://data.theglobalfund.org/location/IDN/grants (accessed Jan 12, 2024).

8 Alegana VA, Okiro EA, Snow RW. Routine data for malaria morbidity estimation in Africa: challenges and prospects. BMC Med 2020; 18: 121.

9 Cotter C, Sturrock HJ, Hsiang MS, et al. The changing epidemiology of malaria elimination: new strategies for new challenges. The Lancet 2013; 382: 900–11.

10 Ferreira MU, Castro MC. Malaria Situation in Latin America and the Caribbean: Residual and Resurgent Transmission and Challenges for Control and Elimination. In: Ariey F, Gay F, Ménard R, eds. Malaria Control and Elimination. New York, NY: Springer, 2019: 57–70.

11 Howes RE, Battle KE, Mendis KN, et al. Global Epidemiology of Plasmodium vivax. Am J Trop Med Hyg 2016; 95: 15–34.

12 Griffin JT, Ferguson NM, Ghani AC. Estimates of the changing age-burden of Plasmodium falciparum malaria disease in sub-Saharan Africa. Nat Commun 2014; 5: 3136.

13 Snow RW, Omumbo JA, Lowe B, et al. Relation between severe malaria morbidity in children and level of Plasmodium falciparum transmission in Africa. Lancet 1997; 349: 1650–4.

14 Sandfort M, Vantaux A, Kim S, et al. Forest malaria in Cambodia: the occupational and spatial clustering of Plasmodium vivax and Plasmodium falciparum infection risk in a cross-sectional survey in Mondulkiri province, Cambodia. Malar J 2020; 19: 413.

15. Subdirektorat Malaria Kementerian Kesehatan Republik Indonesia. Petunjuk Teknis Pencatatan dan Pelaporan Program Malaria Menggunakan Aplikasi SISMAL. Jakarta, 2018.

16 Pedersen EJ, Miller DL, Simpson GL, Ross N. Hierarchical generalized additive models in ecology: an introduction with mgcv. PeerJ 2019; 7: e6876.

17 Simpson GL. Modelling Palaeoecological Time Series Using Generalised Additive Models. Front Ecol Evol 2018; 6. https://www.frontiersin.org/article/10.3389/fevo.2018.00149 (accessed Jan 14, 2022).

18. Wood SN. mgcv: Mixed GAM Computation Vehicle with Automatic Smoothness Estimation. 2021; published online Oct 6. https://cran.r-project.org/web/packages/mgcv/index.html.

19 Herdiana H, Cotter C, Coutrier FN, et al. Malaria risk factor assessment using active and passive surveillance data from Aceh Besar, Indonesia, a low endemic, malaria elimination setting with Plasmodium knowlesi, Plasmodium vivax, and Plasmodium falciparum. Malar J 2016; 15: 468.

20. Rahayu N, Suryatinah Y, Kusumaningtyas H. Discovery of Malaria Cases on Forest Workers In Public Health Center, Teluk Kepayang, Tanah Bumbu, South Kalimantan. BIO Web Conf 2020; 20: 01008.

21 Sutanto I, Soebandrio A, Ekawati LL, et al. Tafenoquine co-administered with dihydroartemisinin–piperaquine for the radical cure of Plasmodium vivax malaria (INSPECTOR): a randomised, placebo-controlled, efficacy and safety study. Lancet Infect Dis 2023; 23: 1153–63.

22 Taylor WRJ, Thriemer K, Seidlein L von, et al. Short-course primaquine for the radical cure of Plasmodium vivax malaria: a multicentre, randomised, placebo-controlled non-inferiority trial. The Lancet 2019; 394: 929–38.

23 Douglas NM, Poespoprodjo JR, Patriani D, et al. Unsupervised primaquine for the treatment of Plasmodium vivax malaria relapses in southern Papua: A hospital-based cohort study. PLOS Med 2017; 14: e1002379.

24. Poespoprodjo JR, Burdam FH, Candrawati F, et al. Supervised versus unsupervised primaquine radical cure for the treatment of falciparum and vivax malaria in Papua, Indonesia: a cluster-randomised, controlled, open-label superiority trial. Lancet Infect Dis 2022; 22: 367–76.

25. Rahmalia A, Poespoprodjo JR, Landuwulang CUR, et al. Adherence to 14-day radical cure for Plasmodium vivax malaria in Papua, Indonesia: a mixed-methods study. Malar J 2023; 22: 162.

26. Plasmanto G. Zoonotic malaria threatens indigenous Orang Rimba in Jambi. Ekuatorial. 2022; published online Feb 15. https://www.ekuatorial.com/en/2022/02/zoonotic-malaria-threatens-indigenous-orang-rimba-in-jambi/ (accessed Dec 12, 2022).

27 Ompusunggu S. Malaria hutan di Provinsi Kalimantan Tengah dan Kalimantan Selatan, Indonesia tahun 2013. Indones J Health Ecol 2015; 14: 145–56.

28. Rohmah K. Perkuat Regulasi untuk Percepatan Eliminasi Malaria di Kaltim. East Kalimantan Prov. Off. Website. 2023; published online July 18. https://diskominfo.kaltimprov.go.id/kesehatan/perkuat-regulasi-untuk-percepatan-eliminasi-malaria-di-kaltim (accessed Sept 26, 2023).

29 Sugiarto SR, Baird JK, Singh B, Elyazar I, Davis TME. The history and current epidemiology of malaria in Kalimantan, Indonesia. Malar J 2022; 21: 327.

30 Hetzel MW, Pulford J, Gouda H, Hodge A, Siba P, Mueller I. The Papua New Guinea national malaria control program: primary outcome and impact indicators, 2009-2014. 2014.

31 Hetzel MW, Saweri OP, Kuadima JJ, et al. Papua New Guinea malaria indicator survey 2016-2017: malaria prevention, infection and treatment. 2018.

32 Ahmed R, Poespoprodjo JR, Syafruddin D, et al. Efficacy and safety of intermittent preventive treatment and intermittent screening and treatment versus single screening and treatment with dihydroartemisinin–piperaquine for the control of malaria in pregnancy in Indonesia: a cluster-randomised, open-la. Lancet Infect Dis 2019; 0. DOI:10.1016/S1473-3099(19)30156-2.

33. CSIRO. Zoonotic Malaria and land use in Indonesia. 2023; published online July 16. https://www.csiro.au/en/work-with-us/industries/agriculture/sustainable-food-and-agriculture-systems/understanding-and-navigating-global-change-in-our-food-systems/linking-malaria-and-land-use (accessed July 17, 2023).

34 Moyes CL, Shearer FM, Huang Z, et al. Predicting the geographical distributions of the macaque hosts and mosquito vectors of Plasmodium knowlesi malaria in forested and non-forested areas. Parasit Vectors 2016; 9: 242.

35 van de Straat B, Sebayang B, Grigg MJ, et al. Zoonotic malaria transmission and land use change in Southeast Asia: what is known about the vectors. Malar J 2022; 21: 109.

36. Meteorology, Climatology, and Geophysical Agency of Indonesia. Climate Variability of Indonesia. 2022. https://iklim.bmkg.go.id/publikasi-klimat/ftp/brosur/LEAFLETINGGRISB.pdf (accessed July 16, 2023).

37 Bin Said I, Kouakou YI, Omorou R, et al. Systematic review of Plasmodium knowlesi in Indonesia: a risk of emergence in the context of capital relocation to Borneo? Parasit Vectors 2022; 15: 258.

38 Barber BE, William T, Grigg MJ, Yeo TW, Anstey NM. Limitations of microscopy to differentiate Plasmodium species in a region co-endemic for Plasmodium falciparum, Plasmodium vivax and Plasmodium knowlesi. Malar J 2013; 12: 8.

39 Coutrier FN, Tirta YK, Cotter C, et al. Laboratory challenges of Plasmodium species identification in Aceh Province, Indonesia, a malaria elimination setting with newly discovered P. knowlesi. PLoS Negl Trop Dis 2018; 12: e0006924.

40 Chin AZ, Avoi R, Atil A, et al. Risk factor of plasmodium knowlesi infection in Sabah Borneo Malaysia, 2020: A population-based case-control study. PLOS ONE 2021; 16: e0257104.

