## supplementary methods and figures for "Shifts in Indonesia’s malaria landscape: an analysis of 2010-2019 routine surveillance data"

### Supplementary appendix

#### Population denominator data

District-level population data were obtained from the 2010 and 2020 census results by Statistics Indonesia.[1,2] The population data comprise the total population by 5-year age groups, from people aged 0-5 years to those over 75 years of age. Simple linear regression models were used to interpolate the yearly district-level population counts for each age group in 2011-2019, based on the population data obtained from the 2010 and 2020 Census. An evaluation of the usage of linear interpolations between two censuses in the United States of America has shown that for demographic characteristics, this interpolation performed well at the county level.[3]

#### Generalised additive model for estimating trends of malaria surveillance metrics

We employed GAM with a negative binomial family, to account for likely overdispersion in the distributions of cases and tests, and population counts as the model offset variable to model malaria cases and tests per 1,000 population.

$$Y_{i,t} \sim NB(N_{i,t}\lambda_{i,t}, r)$$

$$log[E(Y_{i,t})] = \beta_0 + \xi_i + log(N_{i,t}) + g(X_{i,t})$$

with $Y_{i,t}$ as either the number of cases or tests in district $i$ at time (month-year) $t$, $N_{i,t}$ as the population counts in district $i$ at time $t$, $\lambda_{i,t}$ as the case or test rate per population in district $i$ at time $t$, $r$ as the overdispersion parameter of the Negative Binomial (NB) distribution, $\beta_0$ as the model intercept, $\xi_i$ as the district-specific random intercept, and $g(X_{2i,t})$ as the spline function for the time covariate $X_{i,t}$. For comparison, cases and tests were also modelled with GAM with a Poisson family distribution.

Metrics and case profiles measured as proportions were modelled using GAMs with a binomial family and logistic link function.

$$Y_{i,t} \sim Binomial(n_{i,t}, \pi_{i,t})$$

$$log[\frac{\pi_{i,t}}{1 - \pi_{i,t}}] = \beta_0 + \xi_i + \beta_{1i}X_{1i} + g(X_{2i,t})$$

with $Y_{i,t}$ as the number of cases in a specific population group (males or ≥ 15 years old) or infections identified with a specific parasite species (in this case, this means that we can track the proportion of infections that are *P. vivax* relative to *P. falciparum* or other malaria parasite species) in district $i$ at time (month-year) $t$, $n_{i,t}$ as the total reported malaria cases in district $i$ at time $t$, and $\pi_{i,t}$ as the proportion of malaria cases in a specific population group (males or ≥ 15 years old) or infections identified with a specific parasite species (*P. vivax*) in district $i$ at time t. When calculating Test Positivity Ratio (TPR), $Y_{i,t}$ is the number of positive malaria tests reported and $n_{i,t}$ is the number of malaria tests reported.

The malaria surveillance metrics and case profiles modelled are shown in **Table 1,** alongside their respective distribution families. To estimate the regional-level trend lines for all metrics, we calculated the weighted average values of all district-level trend lines within a region. The weighting factors used were: 1) district-level population counts for modelled cases and tests; 2) modelled district-level tests for TPR; and 3) a combination of modelled district-level TPR multiplied by modelled tests for proportions of *P. vivax* cases, cases in males, and cases in individuals over 14 years old.

#### Generalised linear model for modelling the relationship between age of malaria cases and endemicity

First, we assumed a model whereby the Annual Parasite Incidence (API) per 1,000 (at the log-scale) alters the mean age of reported malaria cases ($\mu$) on the geometric scale.

$$\mu = b_1 e^{b_2 log(API)} + b_3$$

The age of cases with mean $\mu$ was assumed to be distributed under the Gamma distribution (a flexible and positive definite distribution) function with a shape parameter ($\alpha$) and a rate parameter ($\beta$). Summing up parameters $b_1$+$b_3$ is the average age in the limit towards API per 1,000 = 0 (i.e., elimination). Parameter $b_3$ represents the extent to which that average age is independent of transmission. The remainder ($b_1$) scaled to the power of $log(API)$ using an $e^{b_2}$ parameterisation, which allows equal prior weight on an increasing or decreasing relationship with transmission.

We assumed the standard deviation ($\sigma$) of the Gamma distribution to be a constant (but estimated within the model). The relationship between $\mu$ and $\sigma$ of the Gamma distribution and their parameters, $\alpha$ and $\beta$, are described as follows:

$$\alpha = \mu^2/\sigma^2$$

$$\beta = \mu/\sigma^2$$

The cumulative density of the Gamma distribution of age of cases was then used to calculate the set of probability $\pi$, which is a vector representing proportions of malaria cases in 0-4 ($\pi_1$), 5-9 ($\pi_2$), 10-14 ($\pi_3$), and ≥ 15 years old ($\pi_4$) age brackets at the national level:

$$\pi_1 = \int_0^5 \frac{\beta^\alpha}{Gamma(\alpha)} y^{\alpha-1} e^{-\beta y}, \ \pi_2 = \int_5^{10} \frac{\beta^\alpha}{Gamma(\alpha)} y^{\alpha-1} e^{-\beta y},$$

$$\pi_3 = \int_{10}^{15} \frac{\beta^\alpha}{Gamma(\alpha)} y^{\alpha-1} e^{-\beta y}, \ \pi_4 = \int_{15}^{\infty} \frac{\beta^\alpha}{Gamma(\alpha)} y^{\alpha-1} e^{-\beta y}$$

To reflect the differences in the population age structure across the country, the proportion was further adjusted by the ratio between the proportion of an age group at the region level ($p_{i,reg}$) to the national level proportion of that specific age group ($p_{i,nat}$). For the age groups 0-4, 5-9, 10-14, and ≥ 15 years old, the ratios, $\gamma_i$, were calculated by:

$$\gamma_i = \frac{p_{i,reg}}{p_{i,nat}}$$

The adjusted proportions for each age group, $\pi'_i$, were calculated by:

$$\pi'_i = \frac{\pi_i \times \gamma_i}{\sum_{j=1}^{4} \pi_j \times \gamma_j}$$

The reported malaria cases in each age bracket of 0-4, 5-9, 10-14, and ≥ 15 years old are represented by $Y_1$, $Y_2$, $Y_3$, and $Y_4$, respectively, where $Y_i \sim Poisson(\lambda_i)$. Hence, the total number of cases for all age groups is $n \sim Poisson(\lambda_1 + \lambda_2 + \lambda_3 + \lambda_4)$. The multinomial distribution for $Y = (Y_1, Y_2, Y_3, Y_4)$ given $n$ (and given $\pi_j = \frac{\lambda_j}{\sum_{i=1}^{k} \lambda_i}$) is defined as:

$$Y \sim Multinomial(n, \pi')$$

with priors for other parameters:

$$b_1 \sim Normal(0,10)$$

$$b_2 \sim Normal(0,1)$$

$$b_3 \sim Normal(0,10)$$

$$\sigma \sim Gamma(1.5,0.1)$$

The model fitting process was implemented in a Bayesian framework using the rstan package in R.[4]

#### Additional figures

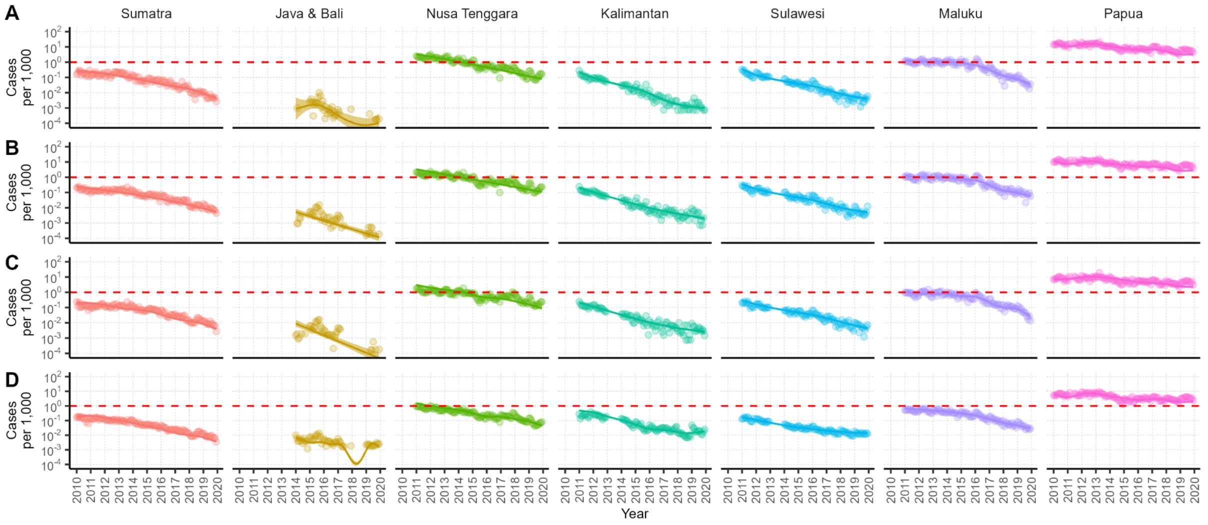

**Figure S1.** Regional-level monthly trends of malaria cases per 1,000 by age group. Solid lines and the shaded areas denote the median and 95% credible intervals of the modelled trends using GAM. The semi-transparent points denote region-level monthly averages from data. Each row denotes modelled estimates for different age groups: **A)** 0-4 years old; **B)** 5-9 years old; **C)** 10-14 years old; and **D)** ≥ 15 years old.

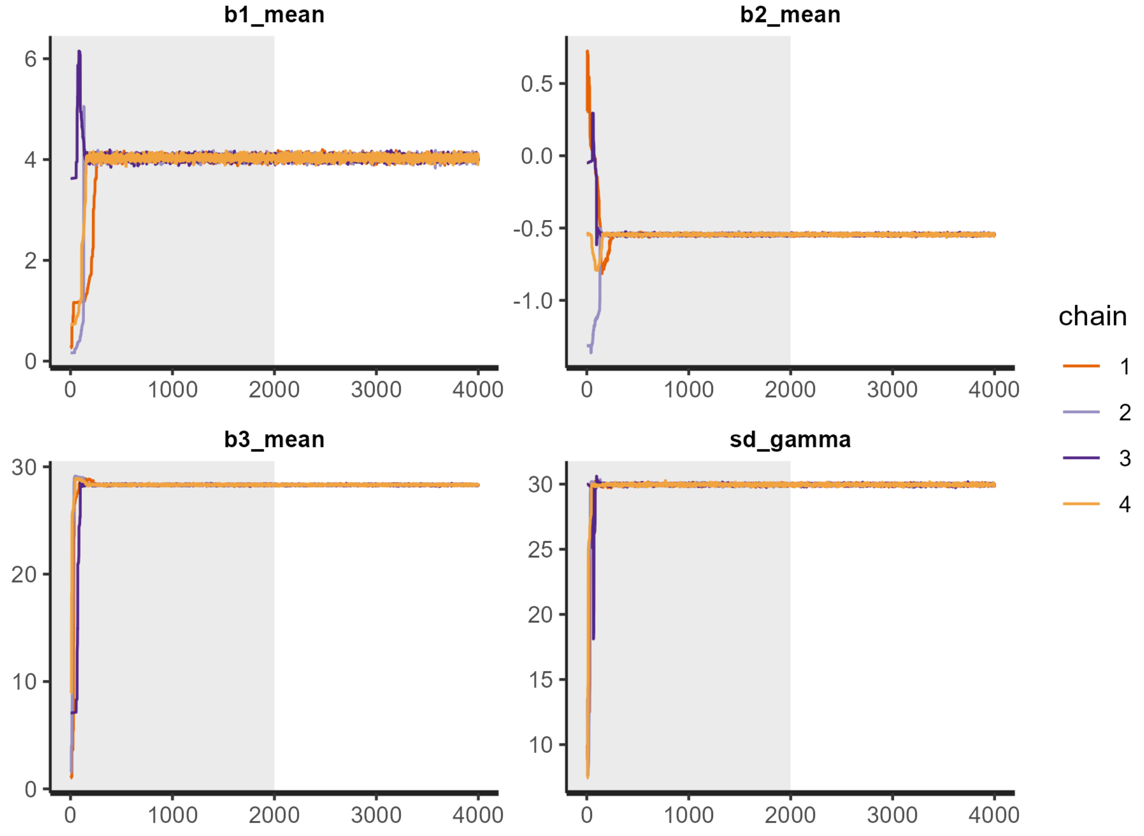

**Figure S2.** Trace plots for GLM fitting in Stan for all fitted model parameters.

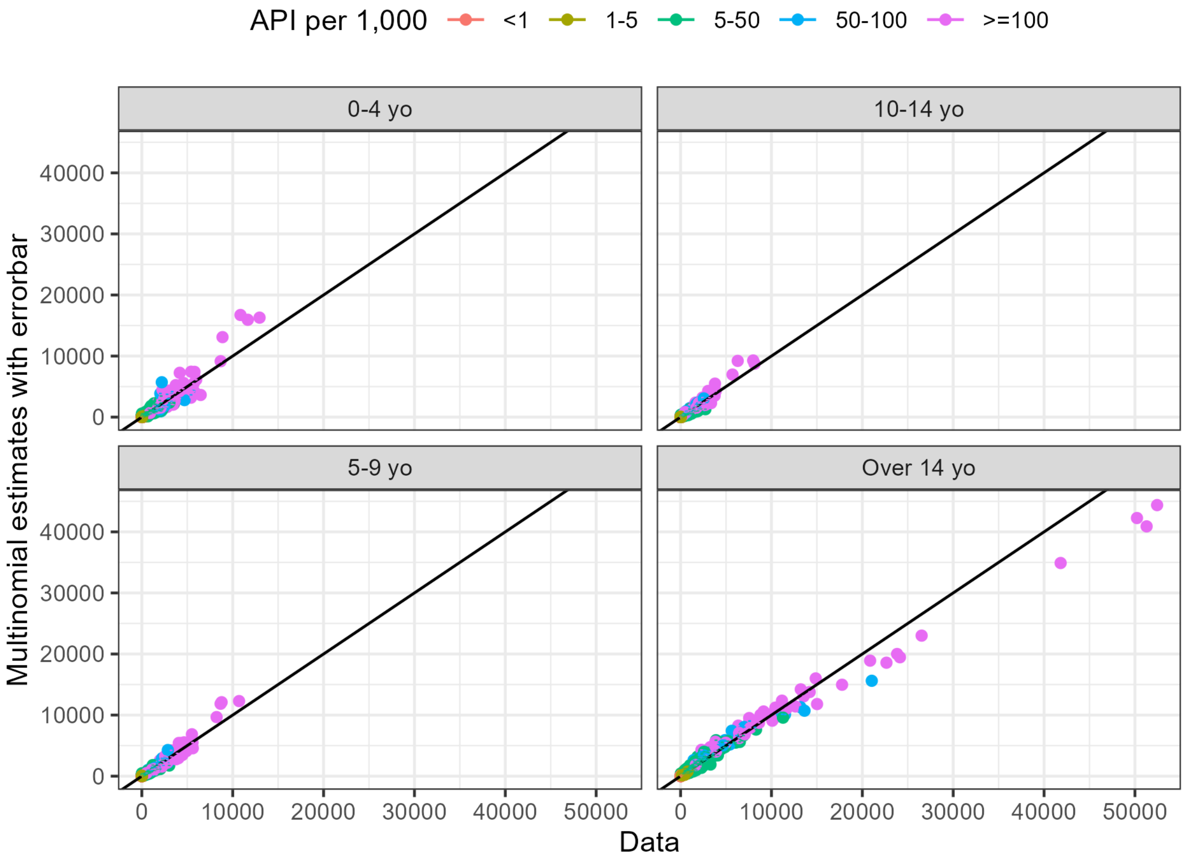

**Figure S3** Plots of case counts by age groups compared to modelled estimates using GLM at the district level. Diagonal lines denote $y = x$.

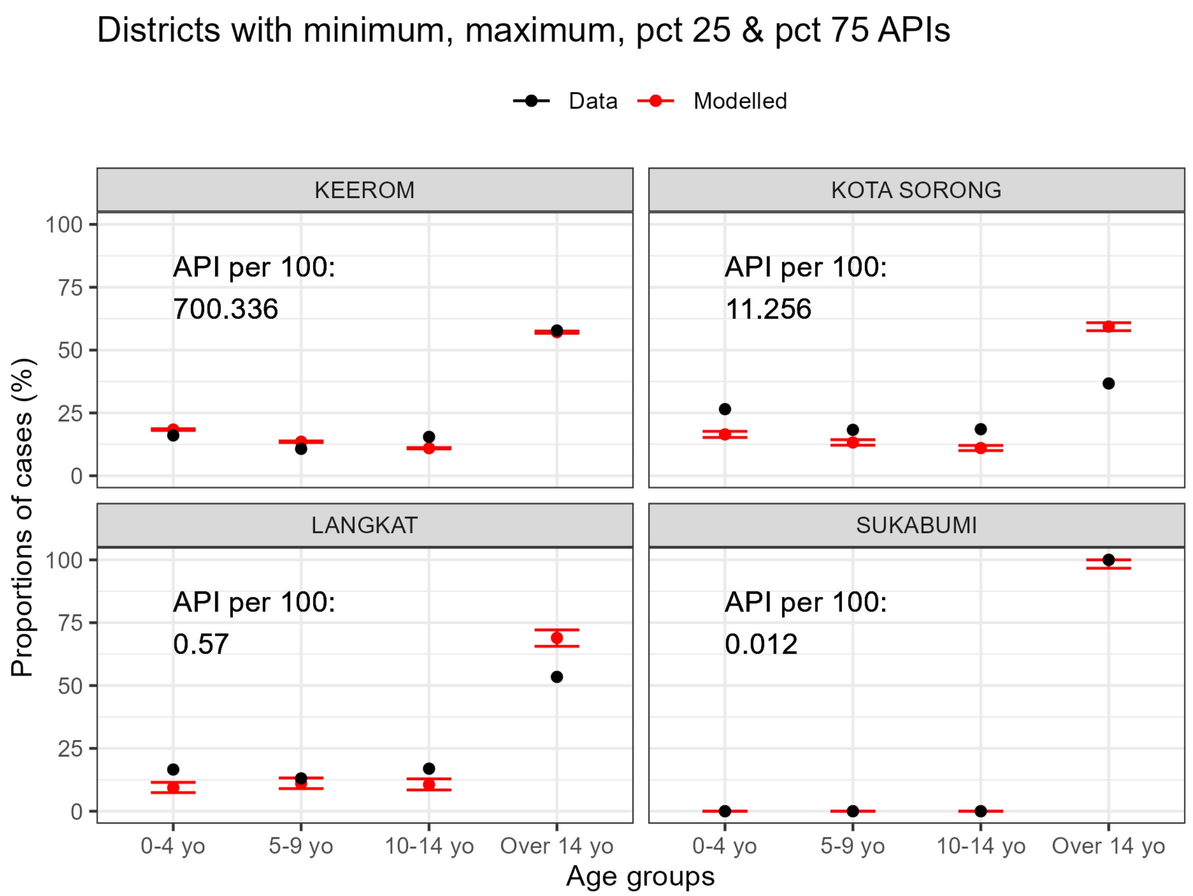

**Figure S4.** Plots of proportions of cases by age groups (black dots) compared to modelled estimates using GLM (red dots with error bars). Data selected were data from districts with the lowest, the highest, 25th percentile, and 75th percentile or the reported API over 2010-2019.

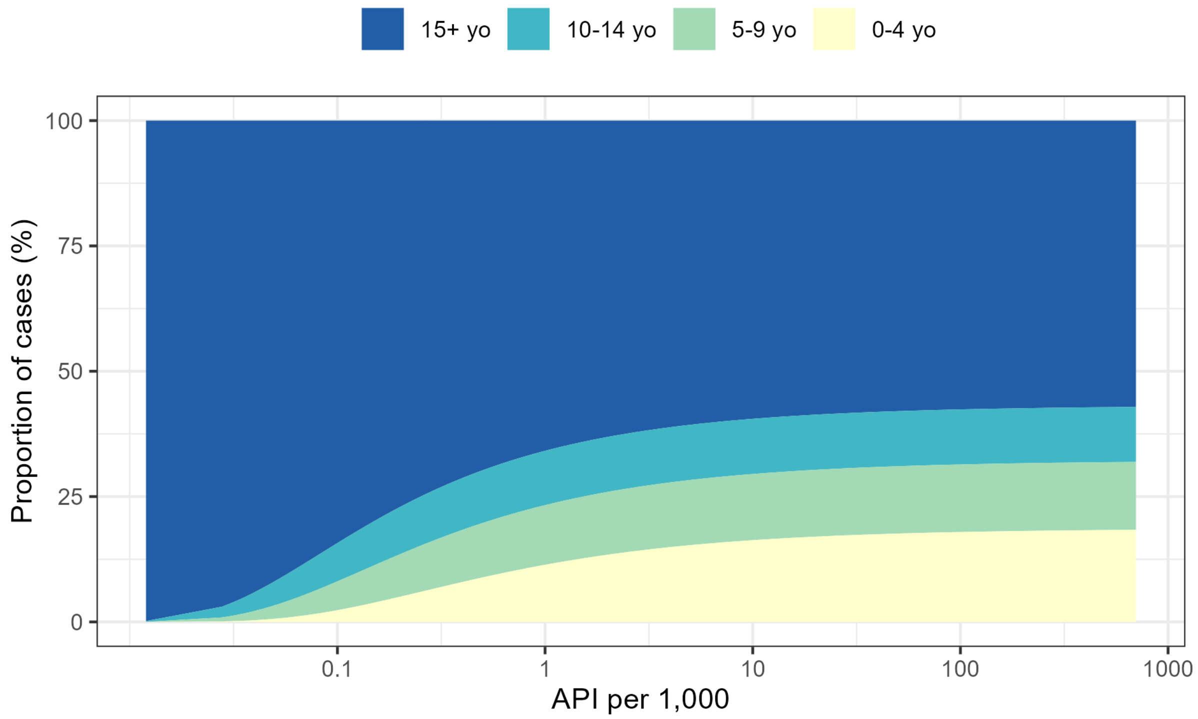

**Figure S5.** Modelled relationship between API per 1,000 and proportion of malaria cases by age group.

**References**

1. Badan Pusat Statistik. Hasil Sensus Penduduk 2010 [Internet]. 2011. Available from: https://sp2010.bps.go.id/
2. Badan Pusat Statistik. Hasil Sensus Penduduk 2020 [Internet]. Jakarta; 2021 Jan [cited 2021 Oct 26]. Available from: https://www.bps.go.id/pressrelease/2021/01/21/1854/hasil-sensus-penduduk-2020.html
3. Weden MM, Peterson CE, Miles JN, Shih RA. Evaluating Linearly Interpolated Intercensal Estimates of Demographic and Socioeconomic Characteristics of U.S. Counties and Census Tracts 2001–2009. Popul Res Policy Rev. 2015 Aug 1;34(4):541–59.
4. Stan Development Team. RStan: the R interface to Stan [Internet]. 2020. Available from: http://mc-stan.org/
